# Gambling in Connecticut Adolescents: Prevalence, Socio-demographic Characteristics, Trauma Exposure, Suicidality, and Other Risk Behaviors

**DOI:** 10.1101/2023.08.11.23294008

**Authors:** Elina A. Stefanovics, Zu Wei Zhai, Marc N. Potenza

## Abstract

Adolescent gambling is a public health concern and has been linked to suicidality and other risk behaviors and poor health correlates. This study examines relationships between adolescents reporting gambling in the past-year and a range of health risk behaviors, traumatic experiences, school performance, and receipt of social support. Data from the 2019 Youth Risk Behavior Survey in Connecticut high-school students stratified by gambling status were examined in bivariate and multivariate analyses. Among 1,807 adolescents, past-year gambling was reported by 453 or 25.4% (95% confidence interval [CI]=22.7-28.1%). Gambling prevalence was higher among older males and lower in adolescents of Asian origin. Gambling was further associated with suicidality and risk behaviors including substance use, smoking (traditional tobacco and electronic vapor use), risky use of digital technologies, unsafe sex, and aggressive behaviors. Gambling was also associated with traumatic experiences, depression/dysphoria, poor academic performance, and less social support from the family and teachers. The results of this study provide an up-to-date estimate of the current prevalence and correlates of gambling among Connecticut adolescents and underscores the importance of routine screening and monitoring of gambling behaviors, as well as interventions for other risk behaviors in this population.

## INTRODUCTION

Gambling involves risking something of value (money or other material goods) on an event of an uncertain outcome in the hope of winning something of greater value. Most people consider gambling to be a harmless form of entertainment and an enjoyable form of recreation (1–3). However, in some individuals gambling may escalate to levels that leads to significant gambling-related personal and social problems, with a sizable minority developing gambling disorder (GD). The etiology of GD is complex and multifactorial and includes a combination of biological, psychological, and social vulnerability that vary across individuals (4).

Adolescents appear particularly vulnerable to developing GD. The prevalence of GD among adolescents is higher than among adults (5–7), with 0.2%–12.3% of youth meeting diagnostic criteria for problem gambling worldwide (7). Brain developmental changes during adolescents years have important implications for behavior (8). Cognitive control is not fully developed and hormonal changes may generate emotional lability and greater responsivities to rewards or stress (9). Young people may inadequately assess risk levels of activities such as gambling (10, 11), have limited life experience, and are more vulnerable to peer influence (12), marketing campaigns (13, 14), and gambling advertisements than adults (15).Thus, adolescents may have multiple risk factors compared to adults (16).

Sociodemographic features of adolescents who gamble include older age and male gender, with older adolescent boys tending to gamble more often (Camirand, 2014) and bet higher amounts of money (17). Male adolescents exposed to gambling at early ages appear at greater risk for developing gambling problems in the future (18). Adult gambling habits often arise from patterns developed during childhood and adolescence (19). Other gambling-associated correlates include substance use and mental health concerns (20, 21), suicidality (22), delinquency (23), and trauma exposure (24). Protective factors against gambling problem may include female gender (25), well-being, social support (26) and parental involvement and monitoring (27). The combination of risk and protective factors as well as availability of specific forms of gambling should be considered for prevention and intervention. For example, when gambling opportunities are limited or non-existent, individuals may engage in other risk behaviors as a means of stress/anxiety reduction.

Adolescents have considerable exposure to gambling. New gambling opportunities are emerging and receiving media coverage, and adolescents participate in multiple types of gambling. Despite age restrictions (Emond & Griffiths, 2020), many youth gamble through lotteries (28), the internet (29), video games (30), slot machines (31), poker (32), and casino games (33), among other forms. High consumption of video games and esports have been linked to gambling, particularly among male youth (34, 35). The practice of competitive video gaming, also known as eSports (e.g., sport competition using video games as platforms for competition between two or more persons (36), sometimes organized into leagues and tournaments (37), is rapidly becoming popular in public schools across the country (38–40). Connecticut was the first state to officially welcome eSports into high schools in 2017. Although gambling can significantly impact adolescents’ lives, it is often unnoticed by teachers and parents until it reaches problematic level (41).

Consistent with the problem behavior theory (PBT), gambling often co-occurs with multiple other risk behaviors (42). According to the PBT, both internal and external factors may contribute to engagement in problematic behaviors (43). The conceptual structure that explains problem behavior consists of three elements: perceived environments (e.g., parental support, encouragement, and control, peer influences), personality (e.g., low self-esteem, low academic achievement, life satisfaction), and behavior (e.g., experimental, or regular use of substances, risky sexual activity, irresponsible driving). There are factors within each element that may be responsible for the balance between risk and protective factors that determines whether or not the individual will engage in problematic behaviors. The PBT also suggests that as one type of problematic behavior increases, the likelihood of the occurrence of other problem behaviors also increases. Co-existence of these behaviors may also be explained by common underlying risk factors, supporting the notion of a problem behavior syndrome (44).

Although the PBT suggests that one risk behavior may elevate the likelihood of others, most studies have not compared a full range of potentially co-occurring problem behaviors in the same analysis, particularly as new emerging risk behaviors related to digital technology use and electronic delivery of drugs like nicotine have become more prevalent. The present investigation seeks to address gaps in the existing research and uses recently collected data to examine the associations between past-year gambling and other health risk behaviors, including substance use, tobacco and electronic vapor product use, sexual behaviors, risky use of digital technologies, and risk activities on school property. To do so, we interrogated data from the year prior to the COVID-19 pandemic. Specifically, in the current study, we sought to provide insight into adolescent gambling and risk behaviors in a contemporary, representative sample of Connecticut high-school students participating in the 2019 Youth Risk Behavior Survey (YRBS). In this exploratory study, we had three specific aims: (1) characterize the prevalence of past-year gambling among Connecticut high-school students; (2) identify sociodemographic characteristics associated with gambling; and (3) assess trauma exposure, other risk behaviors, suicidality, homelessness, health status, academics, and social support associated with gambling. We hypothesized that adolescent gambling would be prevalent, particularly among older males, and would associate with trauma exposure, multiple risk behaviors, and other concerns including suicidality.

## MATERIALS AND METHODS

### Participants

Cross-sectional YRBS data collected in 2019 from public high-school students in Connecticut were analyzed. Altogether, 2,015 students from a total of 33 public, charter, and vocational high schools in Connecticut were surveyed during the spring of 2019. Students completed a self-administered, anonymous, 99-item questionnaire collecting data about demographics characteristics and participation in risk behaviors including ones potentially contributing to unintentional injuries and violence, sexual behaviors potentially contributing to unintended pregnancies and sexually transmitted diseases, alcohol, tobacco and other drug use, and poor physical activity. A description of data collection procedures in the YRBS can be obtained elsewhere (http://www.cdc.gov/yrbss).

The survey procedures have been previously described (45). Briefly, the school response rate was 66%, the student response rate was 82%, and the overall response rate was 54%. The results were representative of all students in grades 9-12. Permission for the survey was obtained through school administrations. Initially, school superintendents were notified regarding selection of their schools to participate. Following superintendent approval, permission from school principals was obtained. Before survey administration, parental permission was obtained. Teachers and/or students of selected classrooms could decline participation. Parents were mailed letters outlining the study, and letters directed parents to contact their schools should they wish to decline their child’s/children’s participation. Active consent, in which parents affirmed and authorized permission, was obtained when required. All procedures were performed in accordance with the 1964 Helsinki Declaration and its amendments. Surveys were administered at each school in a single day. Answers were anonymous and confidential, and students were reminded that participation was voluntary. No personal identifying information was collected. School and classroom codes were removed from the final dataset. The Center for Disease Control (CDC) cleaned and coded the dichotomous variables from the raw data in the final data set. Survey procedures were designed to protect privacy of the students by allowing for anonymous and voluntary participation. During survey administration, students completed the self-administered questionnaire during one class period and recorded their responses directly on a computer-scannable booklet. The CDC’s Institutional Review Board approved the protocol for the YRBS. This research has been deemed exempt from Yale University School of Medicine IRB review by the Human Subjects Committee. This protocol has been determined to be exempt under federal regulation 45 CFR 46.101(b)(2). Exempt studies do not require annual IRB review.

### Measures

***Socio-demographics characteristics*** included age (≤15 years, 16-17 years, ≥18 years), sex (female/male), and race/ethnicity (Black, Hispanic/Latino, White and other).

***Gambling*** was assessed with the question, “During the past 12 months, how many times have you gambled on a sports team, gambled when playing cards or a dice game, played one of your state’s lottery games, gambled on the Internet, or bet on a game of personal skill such as pool or a video game?” If respondents gambled one or more times during the past 12 months, they were classified as having gambling; otherwise, they were classified as having non-gambling.

***Trauma variables:*** Adolescents were assessed on trauma-related measures including feeling unsafe at school in the past 30 days, as well as having experienced threat or injury with a weapon at school, physical fighting, dating violence (sexual and/or physical assault by a dating partner), bullying at school, and electronic bullying (bullied through texting and social media) in the past 12 months; and ever having been forced into sexual intercourse in one’s lifetime. Homelessness was measured by the question, “During the past 30 days, did you ever sleep away from your parents or guardians because you were kicked out, ran away, or were abandoned?”

***Suicidality/self-injury:*** Adolescents were assessed on past-year suicide attempts (yes/no), suicidal ideation (yes/no), and non-suicidal self-injury (NSSI) (yes/no).

***Risky use of digital technologies*** was assessed using 3 questions. Adolescents were asked a questions concerning ***video game and computer use*** other than for school purposes for more than 3 hours daily (yes/no) and whether they were ***talking on a cell phone or texting/e-mailing*** while driving in the past 30 days (yes/no).

#### Substance use

***Medication misuse*** included questions about the use of pain medication without a doctor’s prescription (yes/no) and ever having taken over-the-counter medications to get high (yes/no).

***Binge drinking*** was assessed as heavy drinking of alcohol (≥ 4 drinks in a row for females, ≥ 5 drinks in a row for males) one or more times in the past 30 days (yes/no).

***Lifetime actual and synthetic marijuana use*** was assessed with the questions, “During your lifetime, how many times did you use marijuana (synthetic marijuana)?” Responses were coded as 0 in cases of “never” and 1 if other options were chosen.

***Lifetime electronic vapor use*** was dichotomized using the question, “During your lifetime, how many times did you use electronic vapor?” Responses were coded as 0 in case “never” and 1 if other options were chosen.

***Current marijuana and alcohol use*** was assessed using single survey items (“During the past 30 days, how many times did you use marijuana/alcohol?”) and coded dichotomously yes/no, with a “no” response defined as a “never” response.

#### Tobacco and electronic vapor use

***Current tobacco use:*** Two questions concerned the use of any cigarettes or cigars in the past 30 days (yes/no).

***Current electronic vapor use*** was assessed with the questions, “During the past 30 days, did you

use any electronic vapor product?” and “use them on the school property?” (yes/no).

***Risky sexual behavior*** was assessed with two questions: “Have you ever had intercourse with four or more persons?” and “Did you drink alcohol before the last time you had sex?” (yes/no).

#### Aggressive behavior

***Risk activities on school property*** were assessed with two questions assessing having carried a gun, knife, or club (yes/no) or been offered drugs at school (yes/no) during the past 30 days.

***Physical fights*** reflected past-12-month involvement in a physical fight (yes/no).

***Health-related variables:*** Questions included a student’s subjective consideration of general health as good (yes/no) and sleeping 8 hours or more daily (yes/no). Depressive symptoms were assessed with the question, “In the last 12 months, did you feel sad or hopeless almost every day for more than 2 weeks?”

***Academics*** were assessed with two questions investigating involvement in special education programs (yes/no) and grade average (mostly grades of A or B vs. lower).

***Social support variables*** included family support measured by the question, “Do you agree that your family loves you and gives you help and support when you need it?” Teacher support was measured by the question, “Is there at least one teacher or other adult at school that you can talk if you have a concern?” Mental health support was measured by the question, “Do you receive mental health help and support when needed?”

### Data analysis

Analyses proceeded in three steps. First, to summarize variables, descriptive statistics were computed. Second, Bonferroni-corrected χ^2^ and analyses of variance (ANOVAs) were conducted to compare characteristics (e.g., substance use, tobacco use. electronic vaping, risky sexual behavior, risky use of digital technologies, aggressive behaviors, suicidality/self-injury, trauma variables, subjective health status, academics, and social support) between groups (gambling vs. non-gambling). Second, unadjusted and adjusted weighted frequencies of variables were calculated. Third, a series of multivariable binary logistic regression analyses were conducted to assess relationships and independent associations between group membership and substance use, risky behavior, suicidality/self-injury, trauma variables, health status, academics, and social support outcomes. These analyses were adjusted for sociodemographic variables (age, gender, and race/ethnicity). All analyses were performed using SAS 9.4 statistical software (SAS Institute Inc., Cary, North Carolina, U.S.) (46). To make statistically valid inferences from the sample to the study population, we analyzed data considering the sample design and sample-calculated weights. The survey procedures were employed to compute variances that accurately reflected the complex sample design and estimation procedures. Consequently, sample proportions were based on weighted percentages. Statistical significance was evaluated at the 0.05 level.

### Ethics

The high-school survey and procedures were approved by the Yale School of Medicine IRB, and all procedures were approved by the participating high-schools. Passive consent procedures were adopted for parental consent. Parents of students were notified by mail of the survey and that to exclude their child’s participation in the study, they should contact the school or the study team. Parental permission for their child’s participation was implied if they did not make contact with the team or school. Students were informed at the time of survey administration that it was being used for a study, that their participation was fully voluntary, and that they could refuse to fill out the survey if they wished. Those who did not participate in the survey were allowed to do school work while others worked on the survey. Students were also told not to include identifying information on the survey to maintain anonymity. Students were given a pen to fill out the survey. Procedures were in accordance with the Declaration of Helsinki (2013).

## RESULTS

### Descriptive statistics

Data were available for 2,015 individuals. Respondents who had missing data on the gambling question (n=208; 10%) were excluded from analyses. Thus, the total analytic sample consisted of 1,807 individuals, of whom 891 (49.7%) were males. Approximately one-quarter (n= 453; 25.4%) reported gambling one or more times during the past 12 months. Slightly more than one-third of the sample (n=733, 34.3%) was 15 years old or younger. Almost half of the sample included 16- and 17-year-old students (n=855; 45.7%), with the remainder of the sample (n=213; 11.8%) being aged 18 years and older. Slightly more than half of the sample (n=902; 57.7%) was White; 11.6% (n=159) was Black; 5.1% (n=121) was Hispanic; 3.7% (n=84) was Asian; and 21.9% was multiracial or other race (n=525).

The gambling patterns among persons who reported having gambled in the past 12 months are presented in Figure 1. More than half of adolescents reported that they had gambled 1–2 times in the past 12 months. Almost one-fifth reported having gambled 3–9 times in the same period, and 8.4% reported having gambled 40 or more times. The remainder consisted of less than 12% of the total that reported having gambled 10 to 39 times.

**Figure 1.**
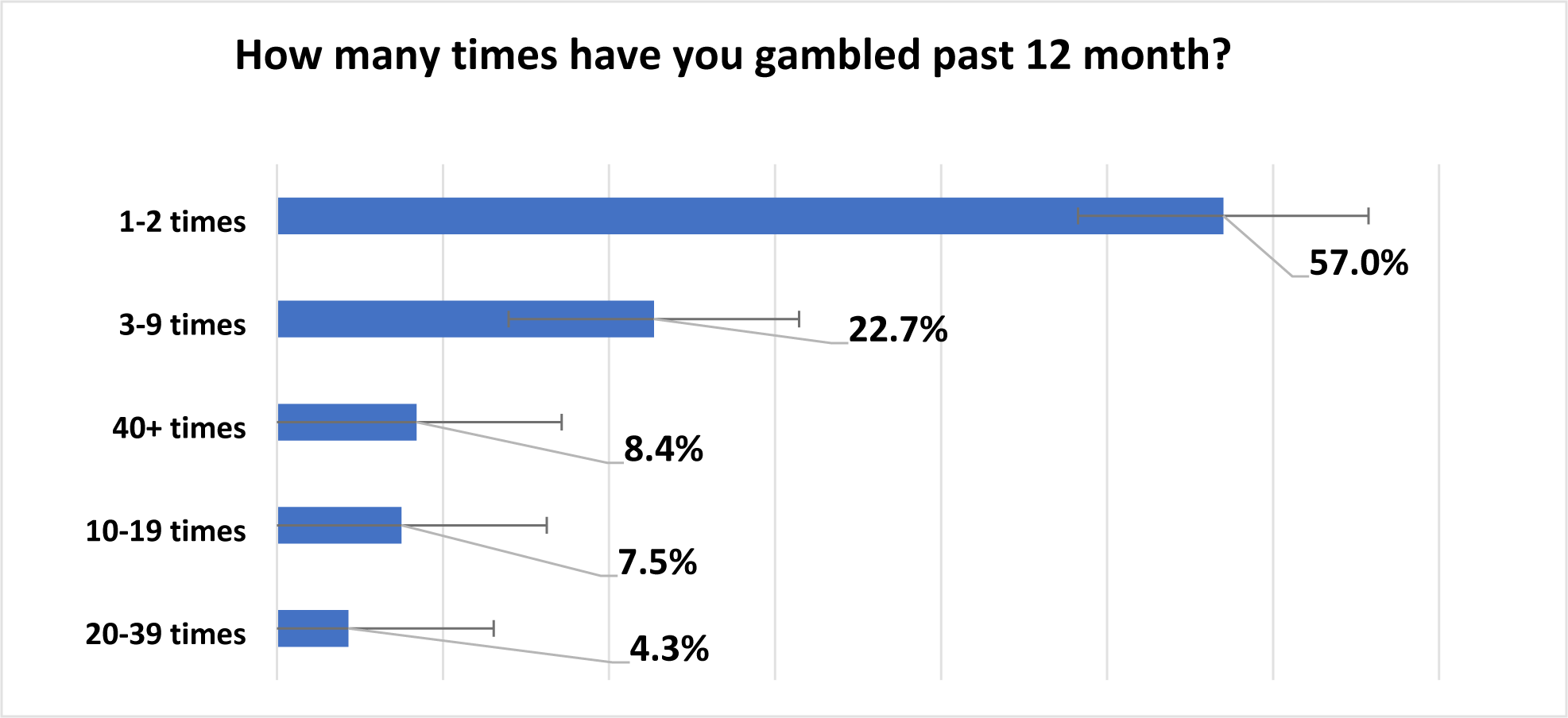
Gambling patterns.

### Bivariate and multivariate analyses

*Table 1* displays the sociodemographic characteristics of adolescents with and without gambling. Adolescents who gambled were more likely to be older and male and less likely to be of Asian heritage.

**Table 1.**
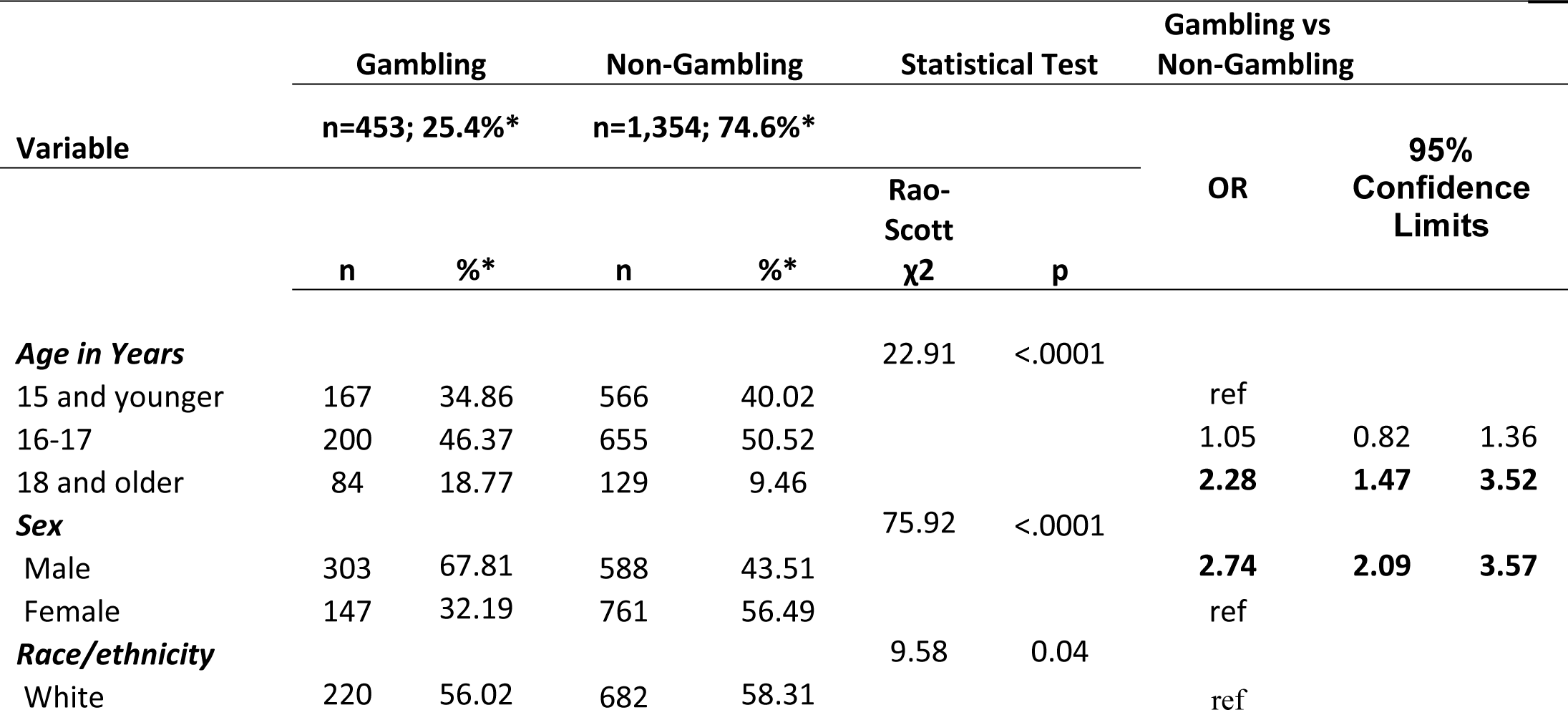
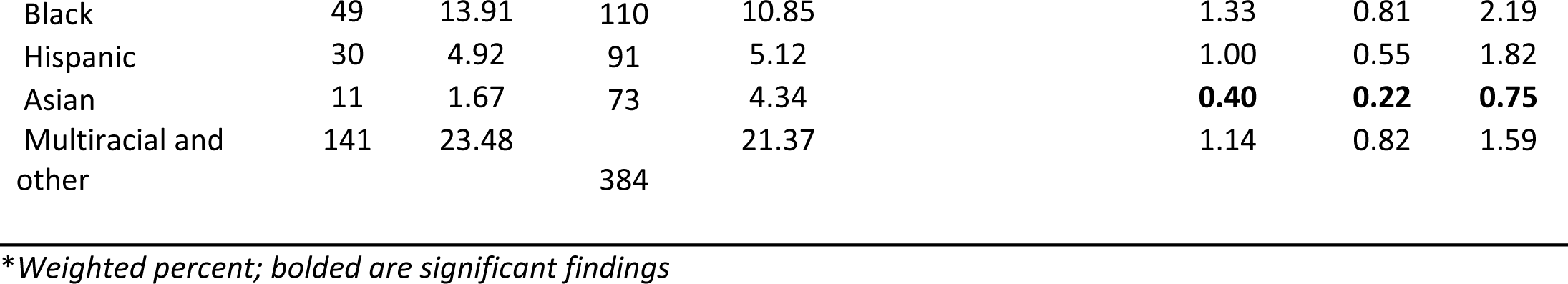
Socio-demographic characteristics of the sample stratified by gambling status (N=1,807)

*Table 2* presents suicidality, traumatic experiences, substance use, and other risk behaviors stratified by gambling status and adjusted for age, sex, and race.

**Table 2.** Traumatic experiences, suicidal/self-injurious and other risk behaviors stratified by gambling status (N=1,807).

|  | Gambling |  | Non-Gambling |  |  |  |  |  |  |
| --- | --- | --- | --- | --- | --- | --- | --- | --- | --- |
| Variables | n=453;<br>25.4%* |  | n=1,354;<br>74.6%* |  | Statistical Test |  | Gambling vs<br>Non-Gambling |  |  |
|  | n | %* | n | %* | Rao-Scott<br>χ2 | p | AOR<br>** | 95%<br>Confidence<br>Limits |  |
| **Traumatic experiences** |  |  |  |  |  |  |  |  |  |
| Bullied at school | 94 | 19.54 | 236 | 17.80 | 0.72 | 0.40 | 1.31 | 0.97 | 1.77 |
| Electronically bullied | 95 | 20.55 | 171 | 12.55 | 14.27 | 0.0002 | **2.15** | **1.61** | **2.89** |
| Homelessness | 68 | 15.48 | 60 | 3.97 | 93.40 | <.0001 | **4.74** | **3.15** | **7.13** |
| Experienced forced sex | 49 | 10.79 | 64 | 4.40 | 30.26 | <.0001 | **3.67** | **2.51** | **5.36** |
| Threatened or injured with weapon<br>at school | 48 | 10.61 | 78 | 5.38 | 8.36 | 0.01 | **2.09** | **1.21** | **3.59** |
| Felt unsafe at school | 54 | 11.15 | 78 | 5.55 | 22.56 | 0.002 | **2.22** | **1.41** | **3.50** |
| Experienced physical or sexual dating<br>violence | 69 | 21.39 | 109 | 14.02 | 5.65 | 0.017 | **2.18** | **1.37** | **3.46** |
| **Substance use** |  |  |  |  |  |  |  |  |  |
| Prescription medications without<br>prescription | 27 | 32.20 | 41 | 28.56 | 0.21 | 0.64 | 1.36 | 0.61 | 3.07 |
| Buy over-the-counter medications to<br>get high | 46 | 10.18 | 37 | 2.47 | 38.60 | <.0001 | **4.07** | **2.55** | **6.49** |
| Binge drinking (PM) | 105 | 24.27 | 124 | 9.42 | 53.67 | <.0001 | **3.30** | **2.49** | **4.37** |
| Alcohol use (PM) | 168 | 38.81 | 279 | 21.59 | 38.72 | <.0001 | **2.53** | **1.89** | **3.39** |
| Marijuana use (PM) | 135 | 30.99 | 250 | 18.70 | 13.72 | 0.0002 | **1.98** | **1.36** | **2.89** |
| Marijuana use (LT) | 207 | 48.82 | 423 | 31.85 | 34.73 | <.0001 | **2.26** | **1.74** | **2.93** |
| Synthetic marijuana use (LT) | 55 | 12.70 | 53 | 3.96 | 32.74 | <.0001 | **3.85** | **2.24** | **6.61** |
| ***Tobacco and electronic vapor use*** |  |  |  |  |  |  |  |  |  |
| Cigarettes use (PM) | 34 | 7.56 | 29 | 2.21 | 23.49 | <.0001 | **3.52** | **1.87** | **6.62** |
| Cigar use (PM) | 47 | 10.75 | 18 | 1.39 | 105.51 | <.0001 | **6.25** | **3.25** | **12.02** |
| Electronic vapor product use (PM) | 155 | 38.24 | 305 | 23.69 | 25.87 | <.0001 | **2.19** | **1.65** | **2.93** |
| Electronic vapor product use (LT) | 253 | 57.79 | 540 | 40.36 | 36.81 | <.0001 | **2.21** | **1.72** | **2.82** |
| Electronic vapor use at school (PM) | 93 | 20.29 | 110 | 8.20 | 36.14 | <.0001 | **2.92** | **2.07** | **4.12** |
| ***Risky Sexual behavior*** |  |  |  |  |  |  |  |  |  |
| Multiple sex partners | 46 | 11.91 | 70 | 5.66 | 15.70 | <.0001 | **1.87** | **1.16** | **3.02** |
| Having sex while drunk | 41 | 31.05 | 49 | 16.75 | 12.11 | 0.0005 | **2.59** | **1.63** | **4.09** |
| ***Risky use of digital technologies*** |  |  |  |  |  |  |  |  |  |
| Play video games or use computer not for school | 402 | 89.43 | 1086 | 80.08 | 12.65 | 0.0004 | **1.73** | **1.12** | **2.68** |
| Talking on cell phone while driving | 119 | 51.97 | 181 | 31.28 | 58.53 | <.0001 | **2.31** | **1.77** | **3.01** |
| Texting/e-mailing while driving | 96 | 45.92 | 143 | 26.79 | 26.79 | 0.0002 | **2.15** | **1.53** | **3.02** |
| ***Suicidality/self-injury*** |  |  |  |  |  |  |  |  |  |
| Attempted suicide | 50 | 10.77 | 71 | 5.14 | 20.11 | <.0001 | **2.82** | **1.87** | **4.26** |
| Considered suicide | 72 | 15.76 | 165 | 11.76 | 3.87 | 0.049 | **1.65** | **1.12** | **2.43** |
| Non-suicidal self-injury (NSSI) | 88 | 17.91 | 207 | 14.55 | 2.86 | 0.01 | **1.55** | **1.14** | **2.12** |
| ***Aggressive behaviors (PM)*** |  |  |  |  |  |  |  |  |  |
| Risk activity on school property (carried a weapon on school property) | 31 | 6.61 | 31 | 2.22 | 31.21 | <.0001 | **2.63** | **1.66** | **4.15** |
| Physical fight | 155 | 34.65 | 201 | 14.19 | 105.25 | <.0001 | **3.09** | **2.32** | **4.11** |

\*weighted %
\*\* AOR adjusted for age, race, and gender; bolded are significant findings
PM=past month
LT=lifetime

Adolescents who gambled reported more traumatic experiences including electronic bullying, forced sex, physical and sexual dating violence, having been threatened or injured with a weapon on school property, having felt unsafe at school, and homelessness. The gambling group was more likely to report alcohol and binge drinking, marijuana and synthetic marijuana use, and use of over-the-counter medication to get high. The gambling group was more likely to report having played video games or using a computer for other than school purposes three or more hours per day, using alcohol and/or marijuana (current and lifetime), engaging in binge drinking, and misusing over-the-counter medication to get high. They were also more likely to report having used tobacco (cigarettes and cigars) and electronic vapor products, been offered drugs at school, engaged in risky sexual behaviors (sex with multiple partners and sex while drunk), and involvement in aggressive behaviors (e.g., carrying a weapon on school property, having been in a physical fight on school property). Adolescents from the gambling group reported they were more likely to have considered suicide and experienced NSSI, and twice as likely to have attempted suicide.

*Table 3* presents subjective health, academic performance, and social support measures.

**Table 3.** Perceived health status, academics and support variables stratified by gambling status (N=1,807)

|  | Gambling |  | Non-Gambling |  |  |  |  |  |  |
| --- | --- | --- | --- | --- | --- | --- | --- | --- | --- |
| Variables | n=453; 25.4%* |  | n=1,354; 74.6%* |  | Statistical Test |  | Gambling vs Non-Gambling |  |  |
|  | n | %* | n | %* | Rao-Scott<br>χ2 | p | AOR*<br>* | 95%<br>Confidence<br>Limits |  |
| **Perceived health status** |  |  |  |  |  |  |  |  |  |
| Dysphoria/depression | 148 | 31.53 | 428 | 30.50 | 0.18 | 0.04 | **1.34** | **1.01** | **1.77** |
| Considered general health as good | 265 | 61.15 | 738 | 55.57 | 5.01 | 0.17 | 1.17 | 0.93 | 1.47 |
| Slept 8 hour or more | 85 | 18.78 | 293 | 21.79 | 1.30 | 0.21 | 0.80 | 0.55 | 1.16 |
| **Academics** |  |  |  |  |  |  |  |  |  |
| Mostly A and B grades | 308 | 68.68 | 1105 | 82.82 | 42.47 | <.0001 | **0.50** | **0.36** | **0.69** |
| Received special education | 51 | 10.16 | 153 | 11.30 | 0.33 | 0.56 | 0.92 | 0.57 | 1.49 |
| **Social Support** |  |  |  |  |  |  |  |  |  |
| Family support | 357 | 79.80 | 1165 | 86.85 | 16.00 | <.0001 | **0.58** | **0.41** | **0.82** |
| Teacher support | 265 | 58.28 | 881 | 65.18 | 6.21 | 0.01 | **0.73** | **0.58** | **0.92** |
| Received mental health help when needed | 65 | 20.98 | 258 | 25.41 | 2.84 | 0.09 | 0.74 | 0.52 | 1.06 |

\*Weighted
\*\*AOR adjusted for age, race, and gender

The gambling group reported poorer school performance, less family and teacher support, and greater likelihood of having felt sad or hopeless almost every day during more than two weeks in the last year.

## DISCUSSION

This study examined 2019 data regarding gambling behaviors and their correlates among high-school students in Connecticut. Approximately one-quarter (25.4%) reported past-year gambling. This estimate appears higher than that previously found (18.6%) among Connecticut adolescents surveyed in 2017 (47). These differences may reflect, in part, the use of slightly different wording in the question to assess gambling involvement in the past year in these two samples. However, other possibilities (e.g., the increasing popularity and social acceptability of sport gambling, especially following the introduction of daily fantasy sports in Connecticut in 2017 and their promotion through frequent advertisements) warrant consideration. The observed gambling prevalence estimate is lower than estimates based on national telephone-based surveys (44.3% to 68%) and in the low range of what was has been reported among students in school-based surveys (24.4% to 86%) and convenience samples (22.5% to 47.4%), although changes over time in the prevalence of adolescent gambling during the early part of the second decade of this millennium should also be considered (48, 49). Although half of the adolescents who have been gambled last year reported doing it only 1 or 2 times, 12.7% of gambling adolescents reported gambling 20 or more times, with 8.4% reporting more than 40 times.

Past-year gambling was higher among male adolescents, consistent with previous findings (7, 18, 50). Previous studies have reported differences in gambling in different racial/ethnic groups (51). Here, adolescents with past-year gambling were less likely to identify as Asian. The extent to which the findings may relate to immigrant status warrants additional examination given that among adults, the prevalence of gambling and problem gambling was markedly lower among first-generation immigrants than among native-born Americans and second and third-generation individuals (52). In an earlier study of Connecticut adolescents, Asian-American adolescents more often reported not gambling (53). However, their study also found that if the Asian-American adolescents did report gambling, they showed higher levels of at-risk/problematic gambling. Taken together, differences in problem-gambling severity and gambling perceptions among Asian youth suggest possible cultural differences (53), with possible differences in parenting styles warranting investigation (54). While currently speculative, this and other possibilities warrant additional investigation.

Consistent with some prior reports, adolescents reporting past-year gambling, relative to those reporting no gambling, were more likely to report past-year suicidal ideation and NSSI and lifetime suicide attempts. One recent study associated gambling and suicide attempts, with the likelihood of suicide attempts increasing with increasing problem-gambling severity in both young women and men (22). Certain groups may be particularly vulnerable. For example, gambling initiation statistically predicted suicidal ideation among Black but not White youth (51). Connecticut high-school students with NSSI have reported more permissive views regarding gambling and were more likely to exhibit at-risk/problem gambling (55). Strong relationships between problem-gambling severity and NSSI suggest that adolescents may engage in gambling as a maladaptive coping strategy to alleviate suffering (55). Taken together, these findings suggest that adolescents who gamble may be at elevated risk for suicidality and NSSI. A history of suicide attempts and suicidal ideation was found to be independently associated with higher rates of risk-taking behavior such as gambling, alcohol and drug use, aggressive behavior, and nicotine use (56). Other studies have also associated health risk behaviors with suicidal ideation and attempts among adolescents, and recognition of these health risk behaviors may be one means of understanding who may be at increased risk of suicidality (57).

Adolescents who gambled reported more frequently substance use, including alcohol, tobacco, and marijuana use, similar to prior observations (21, 48, 58). Such gambling/substance-use relationships were found to extend to use of electronic vaping devices, a newer substance-related concern among youth (59). Further, gambling in adolescents was associated with other risk behaviors including unsafe driving (using cell phone/e-mailing/ texting while driving) and frequent use of computers or cell phones and risky sexual behavior. Gambling was also associated with aggressive behaviors including illegal activities on school grounds and getting into physical fights, as in prior studies of Connecticut high-school students (21). The association between problem-gambling severity and weapon-carrying is also consistent with previous studies of Connecticut high-school students (21, 47).

Traumatic experiences were more prevalent in the gambling group and included bullying, physical violence, forced sex, homelessness, and feelings of being unsafe on school property. Adolescents with past-year gambling also reported low social support. Specifically, they felt less family support and love as well as a lack of support at school, such as a person they could talk to if they had a problem. These findings resonate with earlier findings involving adolescents (60). Youth with gambling problems typically report having experienced more negative life events relative to those who do not gamble (61–63). The extent to which such negative life events may relate to the greater likelihoods for depression/ and poor academic performance observed in this study warrants further examination.

Adolescence is a time when many youths learn how to regulate their emotions and develop personal coping skills for dealing with stress. Some adolescents may deal with stress by engaging in risky behaviors. In such cases, gambling may appear as an attractive means to escape from stressors. According to the Pathways Model (64) and dual process models of addictions that suggest that positive-reinforcements (e.g., gambling to enhance excitement) and negative-reinforcements (e.g., gambling to relieve depressed or stress) may influence participation in and development of habitual gambling in adolescents (65), emotionally vulnerable adolescents under stress may turn to gambling in order to feel important and in control or avoid dealing with emotional problems (66–68). Emotion-based, avoidant, and distraction-orientated coping styles are more often found in adolescents who gamble excessively versus those who do not (69, 70).

Several strengths and limitations of the study are noteworthy. The YRBS is a large epidemiological study involving self-report measures. Self-reported data are subject to biases, such as reliability on memory, social desirability, and honesty of responses. Given the illegal nature of several variables (e.g., gambling among participants younger than 18 years, illicit substance use), it is also possible that participants may have not reported engagement in certain behaviors, leading to overly conservative prevalence estimates. Alternatively, over-reporting of risk behaviors (e.g., for bravado) may also have occurred. Second, the cross-sectional design of this survey study precludes analyses or statements about potential directionality or causality. Third, although this is a representative sample, the survey was conducted in select school districts in one state, and the extent to which our findings generalize to populations outside the state of Connecticut is unclear. Fourth, gambling was assessed with only one question. Thus, types and problem-severity levels of gambling could not be independently determined for analyses. Fifth, many of the questions assessing non-gambling domains were not validated scales. While this may have reduced subject burden, additional studies using empirically validated measures are needed. Sixth, the variables assessed could have been placed in different groups (for example, suicide attempts may have been considered as traumatic, but was grouped with other suicidality measures). Seventh, the COVID-19 pandemic and related stress and combative measures (spatial distancing, closures of schools) may have influenced gambling, other risk behaviors and mental health (71, 72, 73). Thus, additional studies during and following the pandemic are needed.

Overall, results suggest that gambling is a relatively common activity among adolescents. Given that adolescents may be increasingly being exposed to various gambling opportunities online and offline, it is important to identify individuals at risk, monitor the consequences of gambling, and refer individuals for help when needed. This may be particularly relevant in Connecticut and other states in which sports gambling has been recently legalized.

## CONCLUSION

Adolescents who gambled engaged in other risk behaviors, including newer ones that include electronic delivery of substances and use of digital technologies. Given that some of these behaviors occur on school grounds, school administrators and teachers should be aware of them and their implications for adolescent health (74). Intervention and treatment efforts should address multiple domains. Early life stressors, adverse childhood experiences and trauma warrant screening and assessment. Adequate social support may function as a protective factor against gambling and problem gambling. Hence, strengthening adolescents’ social support from school personnel and parents may be beneficial. Social support may be a crucial component of intervention strategies for adolescent gambling and its correlates. Strong familial relationships and open communication between adolescents, parents, and teachers may reduce the likelihood of involvement in risky behaviors such as gambling.

## Data Availability

The Youth Risk Behavior Surveillance System (YRBSS) is a set of surveys that track behaviors that can lead to poor health in students grades 9 through 12. Data are available by site at the national, state, district, territory, and tribal government levels. https://www.cdc.gov/healthyyouth/data/yrbs/contact.htm

https://www.cdc.gov/healthyyouth/data/yrbs/index.htm

